# Understanding multimorbidity trajectories in Scotland: an application of sequence analysis

**DOI:** 10.1101/2022.03.01.22271715

**Authors:** G. Cezard, F. Sullivan, K. Keenan

## Abstract

**BACKGROUND:** Although understanding how multiple conditions develop over time is of growing interest, there is currently little methodological development on the topic, especially in understanding how multimorbidity (the co-existence of at least two chronic conditions) develops longitudinally and in which order diseases occur. Therefore, we aim to describe how a longitudinal method, sequence analysis, can be used to understand the sequencing of common chronic diseases that lead to multimorbidity and the socio-demographic factors and health outcomes associated with typical disease trajectories.

**METHODS:** We use the Scottish Longitudinal Study (SLS) linking the Scottish census 2001 to disease registries, hospitalisation and mortality records. SLS participants aged 40-74 years at baseline were followed over a 10-year period (2001-2011) for the onset of three commonly occurring diseases: diabetes, cardiovascular disease (CVD), and cancer. We focused on participants who transitioned to at least two of these conditions over the follow-up period (N=6,300). We use sequence analysis with optimal matching and hierarchical cluster analysis to understand the process of disease sequencing and to distinguish typical multimorbidity trajectories. Socio-demographic differences between specific disease trajectories were evaluated using multinomial logistic regression. Poisson and Cox regressions were used to assess differences in hospitalisation and mortality outcomes between typical trajectories.

**RESULTS:** Individuals who transitioned to multimorbidity over 10 years were more likely to be older and living in more deprived areas than the rest of the population. We found seven typical trajectories: later fast transition to multimorbidity, CVD start with slow transition to multimorbidity, cancer start with slow transition to multimorbidity, diabetes start with slow transition to multimorbidity, fast transition to both diabetes and CVD, fast transition to multimorbidity and death, fast transition to both cancer and CVD. Those who quickly transitioned to multimorbidity and death were the most vulnerable, typically older, less educated, and more likely to live in more deprived areas. They also experienced higher number of hospitalisations and overnight stays while still alive.

**CONCLUSIONS:** Sequence analysis can strengthen our understanding of typical disease trajectories when considering a few key diseases. This may have implications for more active clinical review of patients beginning quick transition trajectories.

## Background

With increased longevity, our likelihood to develop multiple chronic diseases increases (Divo et al., 2014). Multimorbidity, defined as the co-existence of two or more chronic diseases, has attracted growing scholarly and funders attention in the past two decades (Mercer et al., 2009; The Academy of Medical Science, 2018; Whitty et al., 2020). Multimorbidity is a concern for our ageing societies due to its strong association with higher risk of mortality, higher costs and use of health care services, and worse quality of life (Makovski et al., 2019; Marengoni et al., 2011; Nunes et al., 2016; Wang et al., 2018). However, most multimorbidity research is cross-sectional and evidence about how multimorbidity develops over time remains limited (Head et al., 2021; Marengoni et al., 2011; Xu et al., 2017).

A recent systematic scoping review has highlighted an emerging literature analysing disease trajectories and multimorbidity longitudinally (Cezard et al., 2021). In relation to associative multimorbidity which explores disease clustering (Prados-Torres et al., 2014), the review found only two studies with a longitudinal associative multimorbidity approach (Hsu, 2015; Pugh et al., 2016). The review also identified that while there was an abundance of approaches to study accumulation over time, there was a clear lack of studies focusing on the specific order and sequencing of diseases in multimorbidity trajectories. This points to a lack of methodological approaches to understanding the order of disease onset. Exploring disease order can deepen our understanding of what drives specific trajectories to multimorbidity.

Therefore, our study aims to explore chronic disease trajectories using the concept of sequencing. Sequence analysis, with its origin in molecular biology, is commonly used in the social sciences to study life course patterns and trajectories (Abbott and Tsay, 2000; Brzinsky-Fay and Kohler, 2010; Studer and Ritschard, 2016). Applied to multimorbidity research, sequence analysis can take into consideration the order and the sequence in which diseases occur but also the duration with a first chronic disease before transitioning to another chronic disease i.e. transitioning to multimorbidity. We demonstrate how sequence analysis can help with the visualisation and understanding of the process of transitioning to multimorbidity, using cluster analysis on disease state sequences to distinguish typical disease trajectories. The resulting clusters can be compared according to a range of characteristics to identify, for example, the sociodemographic profile of distinct disease trajectories. Outcomes can also be compared across clusters allowing the identification of specific trajectories at higher risk of worse outcomes.

We provide an example of the value of sequence analysis in researching chronic disease trajectories using three common chronic conditions: diabetes, cardiovascular disease (CVD) and cancer. We explore the link between the social determinants of health and specific multimorbidity trajectories but also whether specific multimorbidity trajectories lead to worse health outcomes. The Scottish Longitudinal Study (SLS) is ideal for such endeavour as it enables the linkage of multiple Scottish censuses (1991-2001-2011), holding individual’s socio-demographic characteristics, to many years of morbidity and mortality records in Scotland. Furthermore, a strong socioeconomic gradient has been demonstrated in the Scottish context, with multimorbidity onset occurring 10-15 years earlier in those living in the most deprived areas compared to those living in the least deprived areas (Barnett et al., 2012).

Therefore, we aim to answer the following research questions:

- What are the typical trajectories to multimorbidity using diabetes, cardiovascular disease, and cancer as exemplars?
- Are there sociodemographic differences in distinct trajectories to multimorbidity based on these three diseases?
- Are specific trajectories to multimorbidity associated with higher health care utilisation and worse outcomes?
- What is the value of sequence analysis in researching multiple chronic disease trajectories?

## Methods

### Data source

The Scottish Longitudinal Study links three Scottish censuses (1991, 2001, and 2011) to a range of administrative data sources including health registers and vital events in a 5.3% sample of the Scottish population (Boyle et al., 2009). Through data linkage, SLS has the advantage to provide socio-demographic determinants from censuses as well as morbidity and mortality information for a representative sample of the population. For our study, the Scottish Census 2001 was linked to hospitalisation, disease registries, exits from Scotland, and mortality records allowing us to investigate the socio-demographic determinants and outcomes of specific disease trajectories in Scotland. We follow SLS participants over a 10 years period, from April 2001 to March 2011. We select participants aged 40-74 years old at the time of the 2001 Census, to focus on understanding disease trajectories from mid-adulthood which can be deemed more preventable than in older ages.

### Disease identification

To accurately determine a sequence of diseases, we first need to identify as precisely as possible the onset of diseases. Our analysis focuses on the onset of three diseases that commonly occur in the population and can be relatively accurately identified from hospitalisation and disease registries. We identified any record of diabetes, CVD, and cancer from hospitalisation data, mental health, diabetes, and cancer registries and with a predefined list of codes from the International Classification of Disease version 10 (ICD10). The list of codes to include in order to define and identify each group of disease can vary. We used the codes E10-E14 to identify a record of diabetes and the codes C00-C97 (excluding non-melanoma skin cancer C44) to identify a record of cancer. We followed the approach of previous Scottish publications using hospitalisation records in Scotland to identify a record of CVD (Fischbacher et al., 2014; Livingstone et al., 2012) and used the codes I20-I25 (ischaemic heart disease), I50 (heart failure), I60-I69 (cerebrovascular diseases), I70 (atherosclerosis) and G45 (transient cerebral ischaemic attacks and related syndromes). Diagnosis records were available from 1997 onwards for diabetes and CVD and from 1980 onwards for cancer. We set any first record of each disease as a first diagnosis and as a proxy for disease onset.

### Sequence creation

Once a first diagnosis for each chronic condition is identified, the date of the first diagnosis is used to order diseases and their co-existence into a sequence of disease states. The resulting sequence is based on the principle of disease combination over time. If we have three diseases A, B, and C, this gives us eight possible states: no disease, A, B, C, AB, AC, BC, and ABC. With four diseases, we would have 16 possible states: no disease, A, B, C, D, AB, AC, AD, BC, BD, CD, ABC, ABD, ACD, BCD, and ABCD. Too many states are difficult to visualise and interpret in sequence analysis. Therefore, we restrict our analysis to multimorbidity trajectories based on three diseases. Note that once a disease is diagnosed, it is kept as present and accumulates with the next disease diagnosed. Consequently, the sequence is set with a number of diseases that can only increase overtime. To account for people that might have left Scotland or died, and thus no longer at risk to develop a new disease that can be identified in Scotland, two states are added: “exit” and “death”. The final set of states included in our sequences is as follows : (1) “no disease”, (2) “diabetes”, (3) “CVD”, (4) “cancer”, (5) “diabetes, CVD”, (6) “diabetes, cancer”, (7) “CVD, cancer”, (8) “diabetes, CVD, cancer”, (9) “exit”, and (10) “death”. Months is used as the time unit for each element of the sequence. Therefore, our sequences are made of 120 consecutive states over a 10-year follow-up period. The first element of the sequence in April 2001 is based on past disease history. For example, if diabetes onset was identified in 2000 and there was no onset of CVD and cancer by the start of the follow-up period, the first state of the sequence is “diabetes”. Subsequent states are constructed by adding up any disease with a first diagnosis up to that month. Once “exit” or “death” occurred, the sequence keeps that state up to the end of the follow-up period (March 2011).

### Covariates

A range of sociodemographic variables were collected in the 2001 Scottish census including age, sex, marital status, household size, and socioeconomic circumstances such as educational level, and household tenure. The Scottish Index for Multiple Deprivation (SIMD), an area-based measure of socioeconomic status created in 2004 is also provided for each SLS participants based on their postcode. We categorise marital status into “single (never married)”, “married”, and “separated, divorced, or widowed”. The household size variable is reduced to three categories: “Household with 1 individual”, “Household with 2 individuals”, and “Household with 3 or more individuals”. Educational level is categorised into “no qualification”, “low qualification” (secondary education and first vocational qualifications) and “high qualification” (higher education, higher vocational and professional qualifications). Household tenure informs on whether individuals lived in a household that they “own”, “private rent”, “social rent” or whether they “live rent free”. SIMD is categorised into quintiles.

### Outcomes

Hospitalisation and mortality data were linked at the individual level for each SLS participants. We choose two hospitalisation outcomes as proxies for health care utilisation: the number of hospitalisations and the number of overnight stays. All-cause mortality was also used as another health outcome. To assess differences between groups, we need to consider a similar period of observation, ensuring a consistent and comparable measure of each outcome across groups. We measure all outcomes from the point of multimorbidity onset (i.e. the point of transition to two chronic diseases) and over a 5-year period.

### Statistical analysis

Sequence analysis is a non-parametric method commonly used in the social sciences to analyse trajectories and social processes. The method has the advantage to provide a holistic view of trajectories, describing how processes evolve over time and when transitions occur. Sequencing (the order of distinct state occurrence), duration (the length of spell in a state) and timing (when transition occurs) are key aspects of a sequence that can be of interest (Studer and Ritschard, 2016). We use single channel sequence analysis with one sequence per person. Sequences show the accumulation and combination the three diseases of interest based on their onset for each individual and as described in the *sequence creation* section. Multiple channel sequence analysis with multiple sequences per person, each sequence describing the trajectory of one disease, is also feasible. However, this approach would not consider sequencing from one disease to another at the individual level but rather concomitant trajectories of each disease. Furthermore, too many channels render interpretation difficult.

First, we describe the sequences using descriptive statistics of the most common reduced sequences, a simplification of sequences focused on sequencing/order of states. For example, for a sequence with the following states “AAABBBCCC”, the associated reduced sequence is “ABC”. Then, we assume that individual trajectories are divided into groups forming typical trajectories. To group sequences together, we need to assess how similar they are. Optimal matching (OM) is the method most often used to assess the dissimilarity between all pairs of sequence (Abbott and Tsay, 2000; Studer and Ritschard, 2016). At the OM stage, choices must be made on costs for three possible operations (substitution, insertion, and deletion) that allows two sequences to match. These costs are set by a substitution matrix (SM) (for substitution operations) and an indel value (for insertion and deletion operations). For this analysis, a SM with a constant value of 1 is chosen with the assumption that “all states are equally different” (cost of 1 for each transition from any state A to B). Alternatives include SM costs based on theory or on a data-driven approach (Studer and Ritschard, 2016). In our preliminary analyses using different forms of SM including the popular data-driven approach with SM based on transition rates, the final clusters obtained were similar to those presented in our results section. A single indel-cost can be determined according to the value we attribute to the aspects of sequencing, duration and timing when assessing similarities between sequences. Since our interest lies mostly in the order of diseases (sequencing) rather than when transition occur (timing), a low indel cost would be appropriate to downplay the cost associated with time lags between two sequences. However, how fast individuals might transition from one state to the other (duration spent in a state) might also be of interest. Therefore, a series of indel values is chosen for sensitivity analyses: 0.5, 1, 1.5, and 5. The OM stage allows us to produce a dissimilarity matrix which can be used in cluster analysis to distinguish typical trajectories. At the cluster analysis stage, we follow a common approach using hierarchical cluster analysis applied to the dissimilarity matrix. Partitioning around the medoid with cluster quality measures (see Additional file 1) is used to decide on the number of clusters with the best clustering. Once clusters are identified, a chronogram (cross-sectional distribution of states at each time t) and a sequence index plot (longitudinal order of states for each individual) are presented to visualise and characterise the typical trajectories represented in each cluster.

In addition, to understand the characteristics associated with typical multimorbidity trajectories, we explore the sociodemographic profile of each trajectory. Descriptive statistics for age, sex, marital status, household size, educational level, household tenure, and SIMD are presented by trajectory cluster. Age and sex-adjusted and multivariate multinomial logistic regressions are also used to understand whether there are significant sociodemographic differences between clusters. We present odd ratios (ORs) and their 95% confidence intervals (CIs).

Finally, we wish to understand whether specific trajectories are associated with greater health care utilisation and worse outcomes. To account for different exposure time per individual, a 5-year denominator is created adjusted for any event of exits or deaths over that period (adjusted person-years). Differences in hospitalisation outcomes (number of hospitalisations and number of overnight stays) are analysed using Poisson regression with an adjusted 5-year person-year as denominator. Risk Ratios (RRs) and their 95% CIs are presented adjusted for sex and age, and then subsequently for the five sociodemographic variables previously described. To account for other comorbidities playing a role in the likelihood of hospitalisations, analyses are further adjusted for a comorbidity count. The comorbidity count is created from 23 Elixhauser comorbidities (excluding eight Elixhauser comorbidities already covered by comorbidities at the core of our analysis i.e. diabetes, CVD, and cancer) and based on the ICD10 codes from the Quan et al. algorithm (Elixhauser et al., 1998; Quan et al., 2005). Cox regression, censored for exit and death, is used to explore cluster differences in 5-year mortality risk. Cox models are also adjusted for age and sex and subsequently for the five sociodemographic variables and the comorbidity count. Hazard Ratios (HRs) and their 95% CIs are presented.

Data preparation, sequence creation, descriptive statistics, and regression analyses were done using SAS version 9.4 (SAS Institute Inc, Cary, NC, USA). Sequence analysis, optimal matching, and cluster analysis were done using the TraMineR and WeightedCluster libraries in R version 3.4.3 (R Core Team, 2017). Graphical representations were created using R.

### Ethics, data access and disclosure

Ethical approval was obtained from the University Teaching and Research Ethics Committee at the University of St Andrews (reference GG14300). This study was also approved by the SLS Research Board (SLS project number 2018_012) and by the Public Benefit and Privacy Panel for Health and Social Care of NHS Scotland (reference 1819-0093). All analyses were performed in accordance with the relevant SLS guidelines and regulations. Data analysis was conducted in a secure environment, the SLS safe haven, at National Records of Scotland, by a named researcher (GC) with appropriate training and clearance. Analyses followed SLS guidelines to ensure the confidentiality of the data. In addition, results were prepared following the SLS statistical disclosure control protocol. Numerators and denominators are presented rounded to the nearest 10 and percentage estimated from rounded numbers. However, model estimates (odd and risk ratios) and their confidence intervals were calculated based on real numbers.

## Results

### Sample characteristics

From the 109,510 participants aged 40 to 74 years old who responded to the Scottish census 2001 (SLS initial sample), we selected the 6,300 participants (6%) who became multimorbid within the next 10 years i.e. transitioned from no or one disease to at least two diseases from a set of three diseases: diabetes, CVD and cancer. Table 1 presents the descriptive statistics for both, the initial sample and subsample, and stratified by sex. At baseline (Scottish census 2001), the subsample is made of individuals who were overall older (62 years old on average vs 55 years old in the SLS initial sample), more likely to be male (58% versus 48%), to have no qualifications (65% versus 47%), to live in more deprived areas (25% versus 19% live in the most deprived quintile), less likely to own their house (63% versus 73%) and slightly less likely to be married (67% versus 69%). Of the individuals in the SLS initial sample, 13% died within the 10-year follow-up period while over a third died in the subsample of individuals who became multimorbid (based on diabetes, CVD and cancer) during that period.

**Table 1.** Socio-demographic profile of the SLS cohort and the sub-cohort.

|  |  | SLS sample<br>SLS participants who responded to the Scottish<br>census 2001, aged 40-74 years old |  |  | Subsample<br>Those for the SLS sample who started with none or<br>one of the three diseases (diabetes, CVD, Cancer)<br>who developed at least two of them within 10<br>years. |  |  |
| --- | --- | --- | --- | --- | --- | --- | --- |
|  |  | Female | Male | Total | Female | Male | Total |
| **Sample size** | **N** | **57,020** | **52,490** | **109,510** | **2,680** | **3,630** | **6,300** |
| Age | Mean (SD) | 55.3 (9.9) | 54.8 (9.7) | 55.1 (9.8) | 62.6 (8.5) | 61.9 (8.4) | 62.2 (8.4) |
| Sex |  |  |  |  |  |  |  |
| Male | N (%) | - | - | 52,490 (48%) | - | - | 3,630 (58%) |
| Marital status |  |  |  |  |  |  |  |
| Single (never married) | N (%) | 4,590 (8%) | 5,660 (11%) | 10,260 (9%) | 180 (7%) | 280 (8%) | 460 (7%) |
| Married | N (%) | 36,600 (65%) | 37,890 (73%) | 74,510 (69%) | 1,490 (56%) | 2,660 (74%) | 4,150 (67%) |
| Divorced/Separated/Widowed | N (%) | 15,450 (27%) | 8,530 (16%) | 23,990 (22%) | 980 (37%) | 650 (18%) | 1,620 (26%) |
| Household size |  |  |  |  |  |  |  |
| One person | N (%) | 11,220 (20%) | 8,330 (16%) | 19,570 (18%) | 820 (31%) | 620 (17%) | 1,450 (23%) |
| Two people | N (%) | 24,650 (43%) | 22,330 (43%) | 47,000 (43%) | 1,300 (49%) | 2,070 (58%) | 3,380 (54%) |
| Three or more people | N (%) | 20,820 (37%) | 21,370 (41%) | 42,200 (39%) | 540 (20%) | 900 (25%) | 1,440 (23%) |
| Educational attainment |  |  |  |  |  |  |  |
| No qualification | N (%) | 25,390 (48%) | 22,000 (45%) | 47,400 (47%) | 1,590 (69%) | 2,060 (63%) | 3,650 (65%) |
| Low qualification | N (%) | 15,230 (29%) | 14,130 (29%) | 29,370 (29%) | 440 (19%) | 710 (22%) | 1,150 (21%) |
| High qualification | N (%) | 11,970 (23%) | 13,000 (26%) | 24,970 (25%) | 260 (11%) | 520 (16%) | 780 (14%) |
| Household tenure |  |  |  |  |  |  |  |
| Own | N (%) | 40,040 (73%) | 37,560 (74%) | 77,630 (73%) | 1,490 (58%) | 2,310 (66%) | 3,800 (63%) |
| Private rent | N (%) | 1,430 (3%) | 1,710 (3%) | 3,140 (3%) | 60 (2%) | 100 (3%) | 160 (3%) |
| Social rent | N (%) | 12,470 (23%) | 10,450 (21%) | 22,940 (22%) | 900 (35%) | 1,000 (29%) | 1,890 (31%) |
| Live rent free | N (%) | 1,260 (2%) | 1,010 (2%) | 2,270 (2%) | 100 (4%) | 90 (3%) | 190 (3%) |
| Scottish Index for Multiple deprivation |  |  |  |  |  |  |  |
| 1- Most deprived | N (%) | 10,720 (19%) | 9,550 (18%) | 20,280 (19%) | 690 (26%) | 890 (25%) | 1,580 (25%) |
| 2 | N (%) | 11,250 (20%) | 10,060 (19%) | 21,320 (19%) | 690 (26%) | 830 (23%) | 1,520 (24%) |
| 3 | N (%) | 11,420 (20%) | 10,470 (20%) | 21,890 (20%) | 550 (21%) | 740 (20%) | 1,290 (20%) |
| 4 | N (%) | 11,530 (20%) | 10,890 (21%) | 22,420 (20%) | 390 (15%) | 610 (17%) | 1,000 (16%) |
| 5- Least deprived | N (%) | 12,120 (21%) | 11,540 (22%) | 23,660 (22%) | 360 (13%) | 560 (15%) | 920 (15%) |
| Death (within 10 years follow-up) | N (%) | 6,190 (11%) | 7,570 (14%) | 13,760 (13%) | 1,030 (38%) | 1,430 (39%) | 2,460 (39%) |

Source: Scottish Longitudinal Study

### Description of trajectories

We found diverse trajectories of multimorbidity based on three diseases. We first describe start and end states and follow on with the most common sequences of states. At the start of the follow-up period (April 2001), 58% of individuals had none of the three diseases of interest and respectively, 17%, 13% and 12% started with CVD, diabetes, and cancer. At the end of the 10-year period (March 2011), 32% had diabetes and CVD, 9% had diabetes and cancer, 16% had CVD and cancer, 4% had complex multimorbidity (diabetes, CVD, and cancer) and 39% had died.

Table 2 shows the 15 most common reduced sequences (orders of diseases) in the subsample. The most common trajectories are those with a transition from no disease to CVD and then to diabetes and CVD (9%) or from no disease to diabetes and then diabetes and CVD (8%) or alternatively directly starting with either CVD or diabetes in 2001 prior transitioning to both (respectively 7% and 6%). The most commonly occurring trajectories beside this first group of four trajectories are a series of trajectories characterised by a transition from CVD to both CVD and cancer, either starting with no disease and later on transitioning to death (6%), or starting directly with CVD and later on transitioning to death (5%), or starting with no disease and with no transition to death in the studied time frame (5%). Then, we find a series of trajectories including transition from cancer to cancer and CVD (4% starting directly with cancer, 4% starting with no disease, 3% starting with no disease and later transitioning to death). About 3% of trajectories have a direct transition from no disease to both diabetes and CVD. Transitions from either diabetes or cancer to both diabetes and cancer are less common.

**Table 2.** List of 15 most common reduced sequences.

| **Disease order** | **Frequency** | **Percentage** |
| --- | --- | --- |
| No -> CVD -> diabetes + CVD | 540 | 9% |
| No -> diabetes -> diabetes + CVD | 500 | 8% |
| CVD -> diabetes + CVD | 420 | 7% |
| Diabetes -> diabetes + CVD | 360 | 6% |
| No -> CVD -> CVD + cancer -> death | 350 | 6% |
| CVD -> CVD + cancer -> death | 310 | 5% |
| No -> CVD -> CVD + cancer | 290 | 5% |
| Cancer -> CVD + cancer | 260 | 4% |
| No -> cancer -> CVD + cancer | 230 | 4% |
| No -> cancer -> CVD + cancer -> death | 220 | 3% |
| No -> diabetes + CVD | 210 | 3% |
| Cancer -> CVD + cancer -> death | 200 | 3% |
| CVD -> CVD + cancer | 190 | 3% |
| No -> diabetes -> diabetes + cancer | 170 | 3% |
| Diabetes -> diabetes + CVD -> death | 170 | 3% |
| All others | 1,880 | 30% |
| **TOTAL** | **6,300** | **100%** |

Source: Scottish Longitudinal Study

### Trajectory (dis)similarities and clustering

To reduce the dimensionality of the variety of disease sequences and identify typical disease trajectories, we first need to compare all pairs of sequences according to their similarities (see the Methods section). Considering the speed of transition from one disease to multimorbidity as an important element in the calculation of dissimilarity between sequences, we choose an indel value of 1.5 for our analysis, slightly higher than 1, the cost of all substitutions in the flat substitution cost matrix. The corresponding cluster quality measures point to a 7-cluster solution (<u>Additional file 1</u>). Note that sensitivity analyses, using different indel cost values (0.5, 1 and 5) at OM stage, point to a best number of clusters between 6 and 8 depending on the indel cost chosen, but all analyses concur to provide reasonable quality measures for a 7-cluster solution (<u>Additional file 1</u>).

Therefore, cluster analysis allows us to differentiate seven typical disease trajectories. These clusters can be visualised using a chronogram (Figure 1) and an index plot (Figure 2). The chronogram shows the distribution of individuals in each state at each time point (cross-sectional) and give us an idea of what state is dominant in each cluster. For example, clusters 3 and 4 are dominated by respectively cancer and diabetes for many years prior to transition to multimorbidity while clusters 5 and 7 show trajectories dominated by multimorbid states, respectively “diabetes, CVD” and “CVD, cancer”. The sequence index plot shows one line per individual’s trajectory (longitudinal). For example, in cluster 5, most individuals start with either diabetes or CVD and quickly transition to having both diabetes and CVD. Graphs combined show that cluster 1 is dominated by individuals starting with none of the three diseases, who transition later to having one disease and usually quickly after a second disease. Therefore, it shows a later and quick transition to multimorbidity within the 10-year period of interest. Cluster 2 is dominated by the “CVD” state for many years prior transitioning to “CVD, diabetes” and to a lesser extent “CVD, cancer”. Cluster 3 shows “cancer” as the dominant state with a later transition to multimorbidity (either “cancer, diabetes” or “cancer, CVD”) and in some case, a transition to complex multimorbidity (“cancer, diabetes, CVD”). Cluster 4 shows a strong dominance of the “diabetes” state with a transition to a second disease after many years in this state. Cluster 5 is dominated by the multimorbid state “diabetes, CVD”. Cluster 6 shows trajectories of individuals who quickly transition to multimorbidity and then death. Cluster 7 is dominated by the multimorbid state “CVD, cancer”. Based on these graphs, we can characterise and label the seven typical disease trajectories as follows: later fast transition to multimorbidity (cluster 1), CVD start with slow transition to multimorbidity (cluster 2), cancer start with slow transition to multimorbidity (cluster 3), diabetes start with slow transition to multimorbidity (cluster 4), fast transition to both diabetes and CVD (cluster 5), fast transition to multimorbidity and death (cluster 6), fast transition to both cancer and CVD (cluster 7).

**Figure 1.**
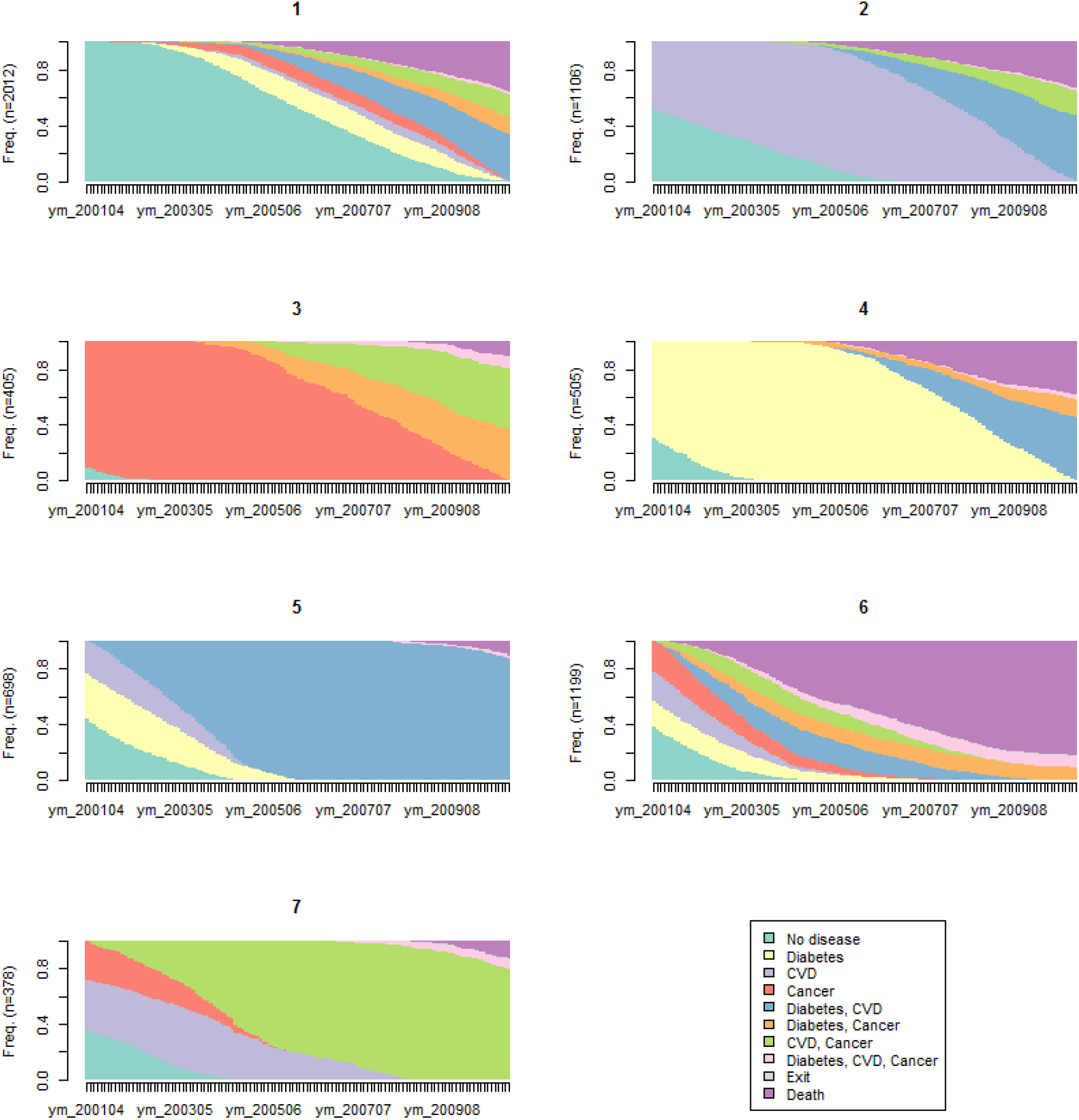
Sequence index plot for the 7-clusters solution. Source: Scottish Longitudinal Study

**Figure 2:**
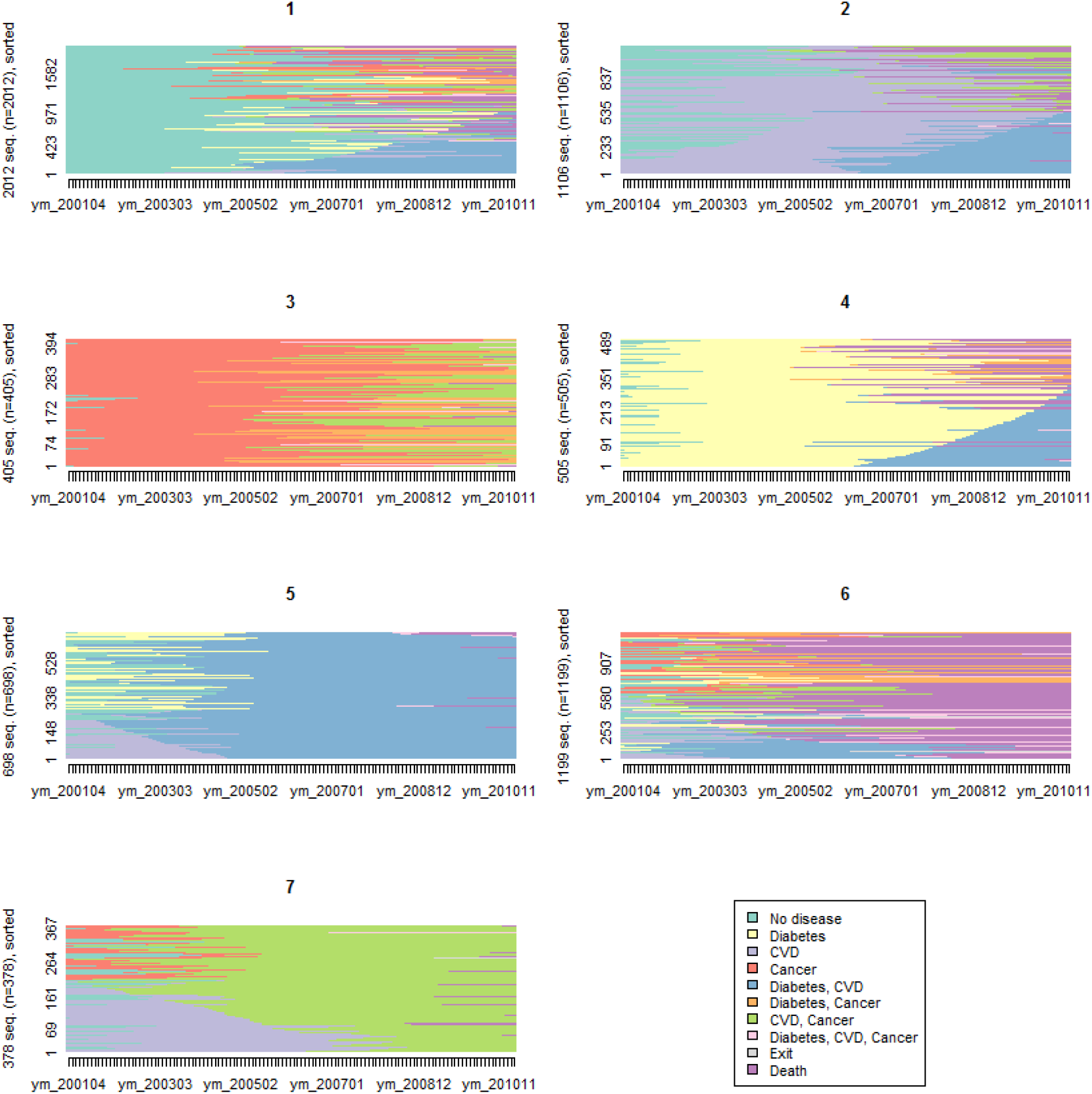
Sequence index plot for the 7-clusters solution. Source: Scottish Longitudinal Study

### Sociodemographic profile of typical trajectories

The descriptive analysis of each cluster (Table 3) shows that cluster 6 (*fast transition to multimorbidity and death*) has the oldest population (65 years old on average), the lowest proportion of married individuals (61%), the highest number of individuals living in a one-person household (27%), the highest proportion of individuals with no qualification (69%), and the lowest proportion of people living in owned properties (58%). By contrast, cluster 3 (*cancer start, slow transition to multimorbidity*) has the highest proportion of individuals living in the least deprived SIMD quintile (20%) and of individuals with higher educational level (19%). Cluster 2 (*CVD start, slow transition to multimorbidity*) has the largest proportion of men (63%).

**Table 3.** Socio-demographic profile of the seven typical disease trajectory clusters.

|  |  | Cluster 1<br>Later<br>fast MM | Cluster 2<br>CVD start<br>slow MM | Cluster 3<br>Cancer start<br>slow MM | Cluster 4<br>Diabetes start<br>slow MM | Cluster 5<br>Fast MM to CVD<br>and diabetes | Cluster 6<br>Fast MM<br>and death | Cluster 7<br>Fast MM to CVD<br>and cancer |
| --- | --- | --- | --- | --- | --- | --- | --- | --- |
| **Sample size** | **N** | **2,010** | **1,110** | **410** | **510** | **700** | **1,200** | **380** |
| Age | Mean (SD) | 61.2 (8.8) | 61.9 (8.3) | 62.6 (8.1) | 61.5 (8.9) | 60.6 (8.6) | 65.0 (7.4) | 63.6 (7.5) |
| Sex |  |  |  |  |  |  |  |  |
| Male | N (%) | 1,150 (57%) | 700 (63%) | 190 (46%) | 290 (57%) | 410 (59%) | 690 (58%) | 200 (53%) |
| Marital status |  |  |  |  |  |  |  |  |
| Single (never married) | N (%) | 160 (8%) | 70 (6%) | 20 (5%) | 50 (10%) | 50 (7%) | 100 (8%) | 20 (5%) |
| Married | N (%) | 1,330 (67%) | 740 (68%) | 280 (70%) | 320 (64%) | 490 (70%) | 730 (61%) | 260 (70%) |
| Divorced/Separated/Widowed | N (%) | 510 (26%) | 280 (26%) | 100 (25%) | 130 (26%) | 160 (23%) | 360 (30%) | 90 (24%) |
| Household size |  |  |  |  |  |  |  |  |
| One person | N (%) | 460 (23%) | 240 (22%) | 90 (22%) | 130 (26%) | 130 (19%) | 320 (27%) | 80 (22%) |
| Two people | N (%) | 1,050 (52%) | 590 (54%) | 250 (61%) | 240 (48%) | 360 (52%) | 650 (55%) | 230 (62%) |
| Three or more people | N (%) | 500 (25%) | 270 (25%) | 70 (17%) | 130 (26%) | 200 (29%) | 210 (18%) | 60 (16%) |
| Educational attainment |  |  |  |  |  |  |  |  |
| No qualification | N (%) | 1,130 (63%) | 660 (68%) | 220 (59%) | 290 (67%) | 410 (64%) | 720 (69%) | 220 (63%) |
| Low qualification | N (%) | 390 (22%) | 190 (20%) | 80 (22%) | 80 (19%) | 150 (23%) | 200 (19%) | 70 (20%) |
| High qualification | N (%) | 280 (16%) | 120 (12%) | 70 (19%) | 60 (14%) | 80 (13%) | 120 (12%) | 60 (17%) |
| Household tenure |  |  |  |  |  |  |  |  |
| Own | N (%) | 1,250 (65%) | 640 (60%) | 270 (71%) | 310 (65%) | 420 (61%) | 660 (58%) | 250 (68%) |
| Private rent | N (%) | 60 (3%) | 30 (3%) | - | 10 (2%) | 20 (3%) | 30 (3%) | 10 (3%) |
| Social rent | N (%) | 570 (30%) | 350 (33%) | 100 (26%) | 140 (29%) | 230 (33%) | 410 (36%) | 100 (27%) |
| Live rent free | N (%) | 50 (3%) | 40 (4%) | 10 (3%) | 20 (4%) | 20 (3%) | 40 (4%) | 10 (3%) |
| Scottish Index for Multiple deprivation |  |  |  |  |  |  |  |  |
| 1- Most deprived | N (%) | 490 (24%) | 310 (28%) | 70 (17%) | 120 (23%) | 170 (24%) | 340 (28%) | 80 (21%) |
| 2 | N (%) | 470 (23%) | 280 (25%) | 90 (22%) | 130 (25%) | 170 (24%) | 280 (23%) | 100 (26%) |
| 3 | N (%) | 410 (20%) | 210 (19%) | 90 (22%) | 120 (23%) | 160 (23%) | 230 (19%) | 70 (18%) |
| 4 | N (%) | 340 (17%) | 160 (15%) | 80 (20%) | 80 (15%) | 110 (16%) | 180 (15%) | 70 (18%) |
| 5- Least deprived | N (%) | 300 (15%) | 140 (13%) | 80 (20%) | 70 (13%) | 90 (13%) | 170 (14%) | 70 (18%) |
| Death (within 10 years follow-up) | N (%) | 730 (36%) | 370 (33%) | 40 (10%) | 200 (39%) | 80 (11%) | 990 (83%) | 60 (16%) |

Source: Scottish Longitudinal Study

Using multinomial logistic regression, we test whether those sociodemographic differences between clusters are statistically significant. Cluster 6 (*fast transition to multimorbidity and death*) appears to have the worst sociodemographic profile and is therefore taken as the reference (OR=1). Sex and age adjusted multinomial logistic regression (Figure 3 and <u>Additional file 2</u>) confirms that all clusters have a significantly younger population than cluster 6. Individuals in cluster 2 (*CVD start, slow transition to multimorbidity*) are significantly more likely to be males compared to individuals of cluster 6. Individuals of cluster 3 (*cancer start, slow transition to multimorbidity*) are more likely to be females compared to individuals of all other clusters (confidence intervals did not overlap) apart from those of cluster 7 (*fast transition to both cancer and CVD*). Individuals of cluster 6 are the least likely to be married or separated, divorced, or widowed compared to single. However, marital status differences are not necessarily significant. Clusters 3 and 7, both involving multimorbidity trajectories with cancer, have the higher odd ratios of married individuals rather than single. Individuals of clusters 3 (*cancer start, slow transition to multimorbidity*) and 7 (*fast transition to both cancer and CVD*) are more likely to live in a 2-people household and individuals of clusters 1 (*later fast transition to multimorbidity*), 2 (*CVD start, slow transition to multimorbidity*) and 5 (*fast transition to both diabetes and CVD*) are more likely to live in a 3 or more people household versus a 1-person household compared to individuals of cluster 6. Individuals of clusters 1 (*later fast transition to multimorbidity*), 3 (*cancer start, slow transition to multimorbidity*) and 7 (*fast transition to both cancer and CVD*) have a relatively better socioeconomic profile, more likely to have higher educational levels (than no qualification), to live in an owned household (than social rent) and to live in less deprived areas compared to individuals of cluster 6. In multivariate analysis (<u>Additional file 3</u>), some differences previously observed remain significant i.e. individuals of clusters 3 and 7 (trajectories both involving cancer) are more likely to be married than single, individuals of cluster 3 (*cancer start, slow transition to multimorbidity*) are better-off socioeconomically, having higher levels of educational attainment and living in less deprived areas, and individuals of clusters 1 and 7 are more likely to live in an owned household than individuals of cluster 6.

**Figure 3.**
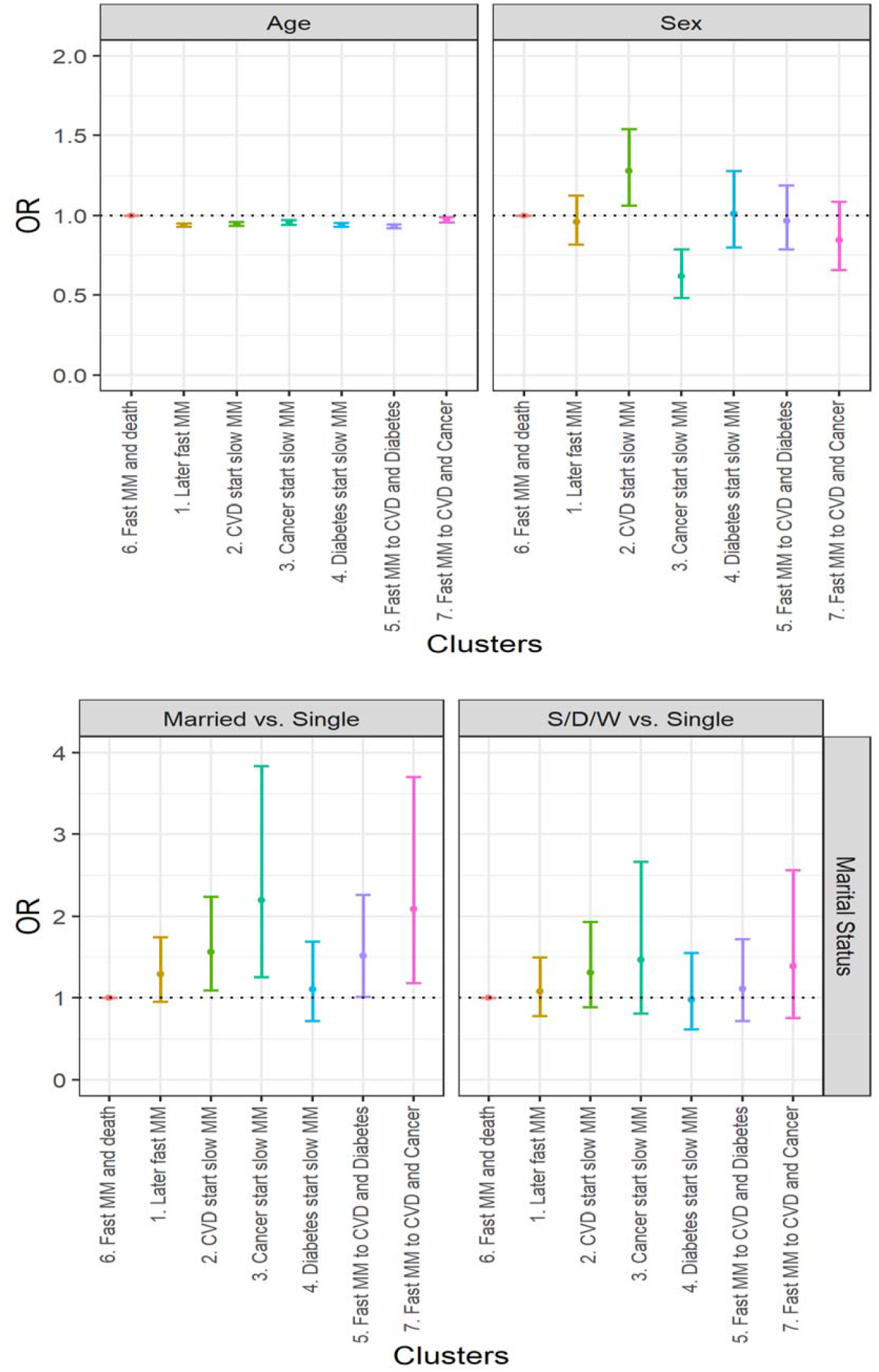
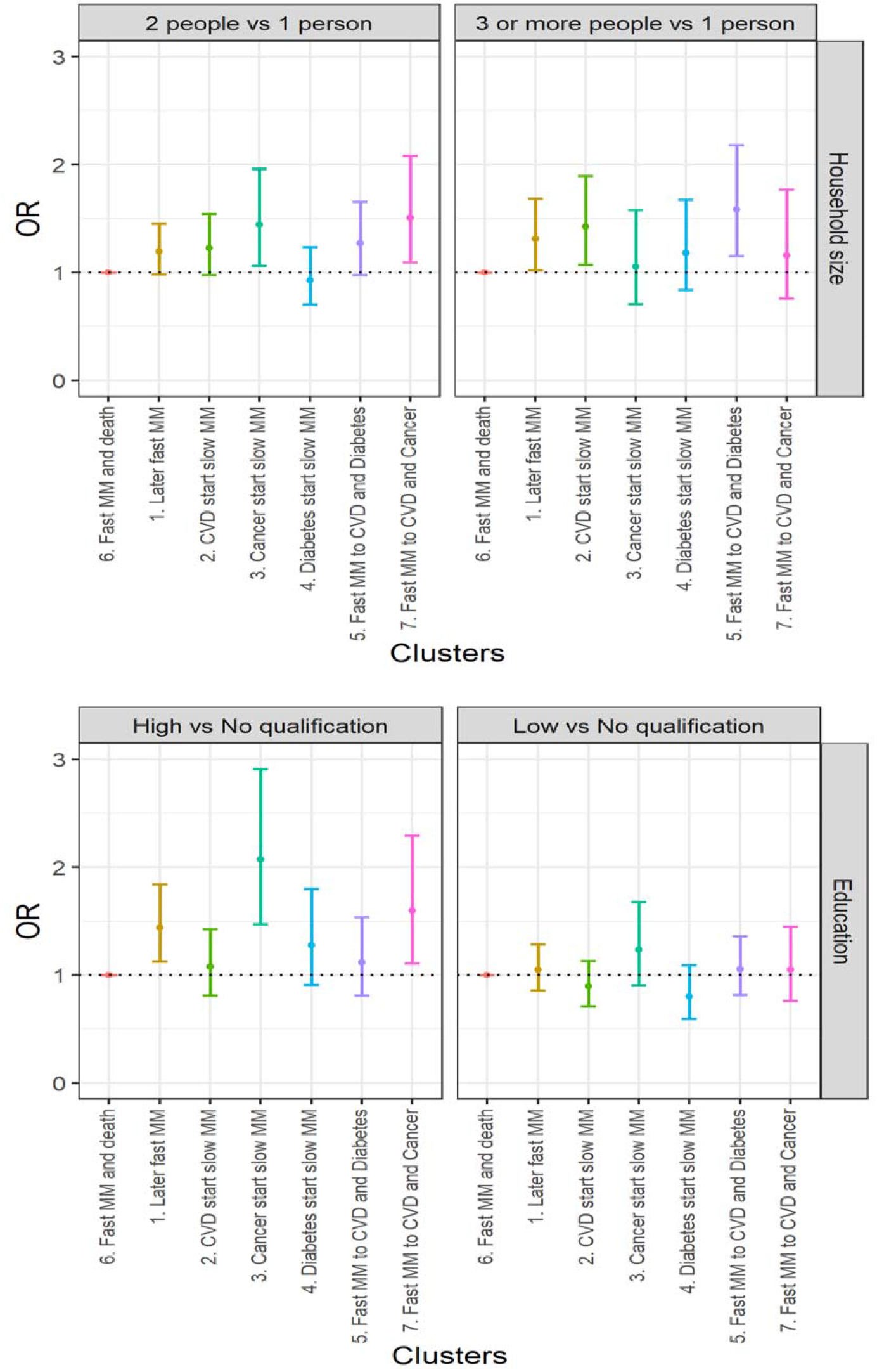
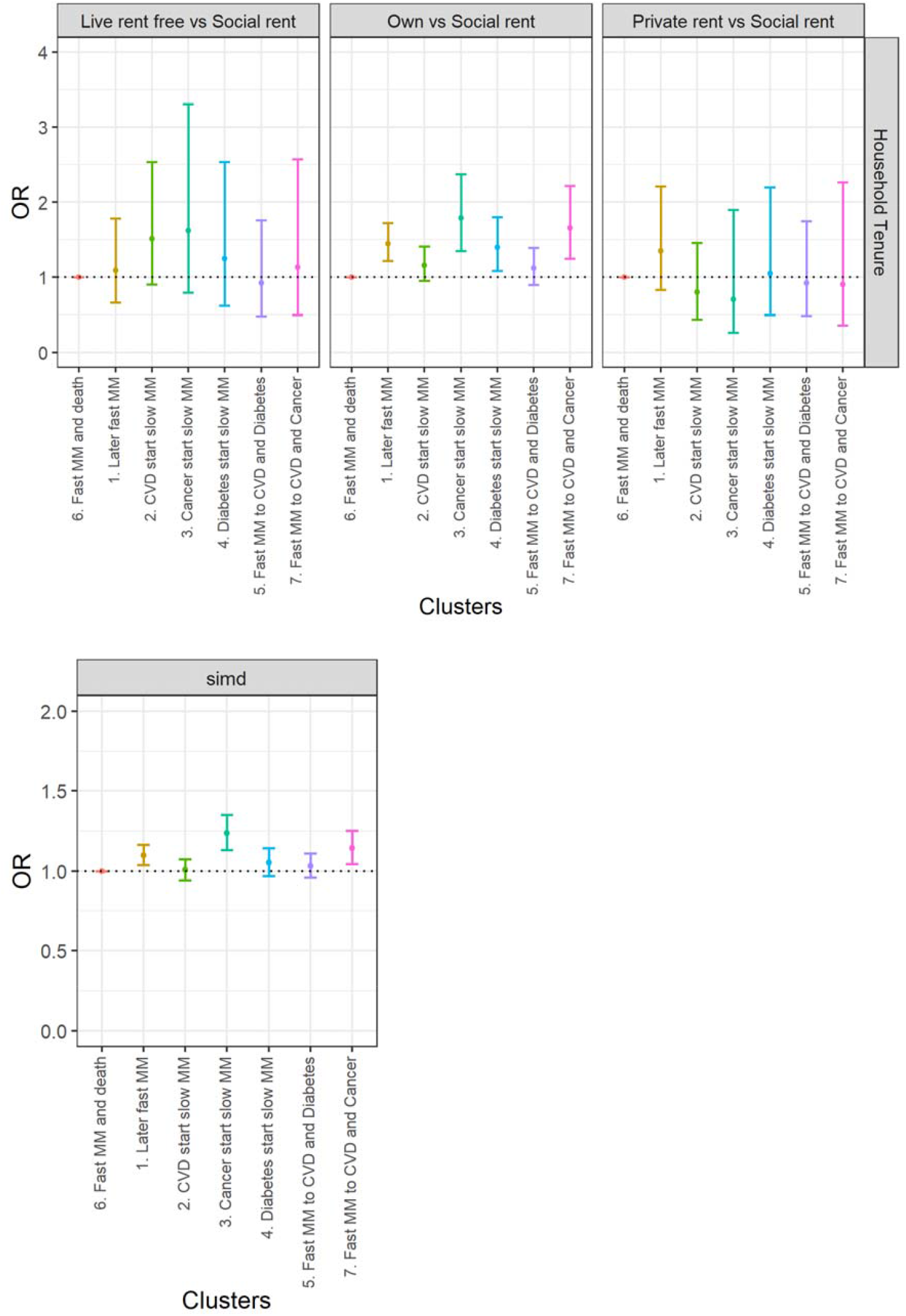
Age- and sex-adjusted socioeconomic differences in typical multimorbidity trajectories. Source: Scottish Longitudinal Study

### Hospitalisation outcomes post multimorbidity onset by cluster

Our subsample of individuals who became multimorbid based on diabetes, cancer and CVD experienced seven hospitalisations on average over a 5-year period post multimorbidity onset. The average number of overnight stays was 42 over the same period. Table 4 shows cluster differences in the number of hospitalisations and overnight stays post multimorbidity onset and over a 5-year period, taking cluster 6 (*fast transition to multimorbidity and death*) as the reference cluster (RR=1). With adjusted person-year at risk for exit and death, sex- and age-adjusted risk ratios shows that individuals of cluster 6 (*fast transition to multimorbidity and death*) have significantly more hospitalisations and overnight stays than individuals of any other cluster. However, this difference in hospitalisation outcomes is not significant for individuals of cluster 4 (*diabetes start, slow transition to multimorbidity*) compared to those of cluster 6. This typical trajectory starting with diabetes and slowly transitioning to multimorbidity (cluster 4) is related to relatively more hospitalisations and overnight stays than most other trajectory types (CIs mostly do not overlap). Individuals of cluster 5 (*fast transition to both diabetes and CVD*) have the lowest risks of hospitalisations within a 5-year period post multimorbidity onset. Further adjusting the sex- and age-adjusted model for the five sociodemographic variables do not change the patterns observed. Additional adjustment for the other comorbidities slightly widens differences between cluster 4 and 6 rendering them significant.

**Table 4.** Cluster differences in risk of hospitalisations and overnight stays following multimorbidity onset.

| Cluster | Number of hospitalisations * | Person-Year ** | Model adjusted for age and sex | Model adjusted for age, sex, marital status, household size, education, household tenure, and SIMD | Model adjusted for age, sex, marital status, household size, education, household tenure, SIMD, and comorbidity count |
| --- | --- | --- | --- | --- | --- |
|  |  |  | Risk of hospitalisation |  |  |
|  |  |  | RR [95% CI] | RR [95% CI] | RR [95% CI] |
| 6 (reference) | 8640 | 2820 | 1 | 1 | 1 |
| 1 | 14290 | 5980 | 0.81 [0.76,0.87] | 0.82 [0.74,0.90] | 0.77 [0.70,0.85] |
| 2 | 6750 | 3370 | 0.68 [0.59,0.78] | 0.67 [0.60,0.76] | 0.65 [0.58,0.73] |
| 3 | 2330 | 1650 | 0.47 [0.43,0.52] | 0.49 [0.41,0.59] | 0.50 [0.42,0.60] |
| 4 | 3940 | 1460 | 0.92 [0.84,1.01] | 0.91 [0.80,1.04] | 0.89 [0.75,0.99] |
| 5 | 3240 | 3070 | 0.36 [0.33,0.39] | 0.36 [0.32,0.41] | 0.36 [0.32,0.40] |
| 7 | 3090 | 1690 | 0.60 [0.52,0.69] | 0.65 [0.56,0.76] | 0.65 [0.56,0.75] |

| Cluster | Number of overnight stays * | Person-Year ** | RR [95% CI] | RR [95% CI] | RR [95% CI] |
| --- | --- | --- | --- | --- | --- |
|  |  |  | Risk of overnight stay |  |  |
|  |  |  | RR [95% CI] | RR [95% CI] | RR [95% CI] |
| 6 (reference) | 60650 | 2820 | 1 | 1 | 1 |
| 1 | 75490 | 5980 | 0.68 [0.63,0.74] | 0.69 [0.59,0.79] | 0.63 [0.55,0.73] |
| 2 | 33710 | 3370 | 0.53 [0.44,0.64] | 0.54 [0.44,0.66] | 0.51 [0.42,0.62] |
| 3 | 9450 | 1650 | 0.28 [0.22,0.36] | 0.29 [0.23,0.36] | 0.30 [0.24,0.37] |
| 4 | 24270 | 1460 | 0.91 [0.82,1.02] | 0.88 [0.73,1.05] | 0.81 [0.67,0.97] |
| 5 | 18350 | 3070 | 0.32 [0.24,0.41] | 0.34 [0.23,0.50] | 0.33 [0.22,0.51] |
| 7 | 15230 | 1690 | 0.43 [0.26,0.70] | 0.43 [0.26,0.71] | 0.43 [0.26,0.71] |

Source: Scottish Longitudinal Study
\* Number of hospitalisations and overnight stays are calculated over 5 years post multimorbidity onset.
\*\* Person-year were calculated over 5 years post multimorbidity onset and adjusted for any exit or death within that period.

### Mortality outcome post multimorbidity onset by cluster

As the typical multimorbidity trajectory of cluster 6 is characterised by a fast transition to death, this cluster is taken as the reference cluster in Cox regression models (HR=1). Table 5 confirms that individuals of all other clusters are significantly less likely to die within 5 years post multimorbidity onset than individuals of cluster 6. However, cluster 6 aside, there are differences between those clusters in relation to their mortality risk. Individuals of cluster 3 (*cancer start, slow transition to multimorbidity*), cluster 5 (*fast transition to both diabetes and CVD*), and cluster 7 (*fast transition to both cancer and CVD*) are significantly less likely to die within 5 years of multimorbidity onset than individuals of clusters 1 (*later fast transition to multimorbidity*), 2 (*CVD start, slow transition to multimorbidity*) and 4 (*diabetes start with slower transition to multimorbidity*) as confidence intervals do not overlap. Additional adjustment for the five sociodemographic variables previously described do not change the trends in mortality risk differences, nor does further adjustment for comorbidity counts.

**Table 5.** Cluster differences in risk of mortality following multimorbidity onset.

| Cluster | Number of deaths * | N | Model adjusted for age and sex | Model adjusted for age, sex, marital status, household size, education, household tenure, and SIMD | Model adjusted for age, sex, marital status, household size, education, household tenure, SIMD, and comorbidity count |
| --- | --- | --- | --- | --- | --- |
|  |  |  | HR [95% CI] | HR [95% CI] | HR [95% CI] |
| 6 (reference) | 1100 | 1200 | 1 | 1 | 1 |
| 1 | 1350 | 2010 | 0.63 [0.58,0.69] | 0.63 [0.57,0.69] | 0.60 [0.54,0.65] |
| 2 | 720 | 1110 | 0.58 [0.52,0.64] | 0.56 [0.50,0.62] | 0.54 [0.48,0.60] |
| 3 | 210 | 410 | 0.32 [0.27,0.37] | 0.33 [0.28,0.39] | 0.33 [0.28,0.40] |
| 4 | 370 | 510 | 0.74 [0.66,0.84] | 0.77 [0.67,0.88] | 0.72 [0.63,0.83] |
| 5 | 370 | 700 | 0.31 [0.27,0.35] | 0.30 [0.26,0.35] | 0.30 [0.26,0.34] |
| 7 | 240 | 380 | 0.34 [0.29,0.39] | 0.35 [0.30,0.41] | 0.35 [0.30,0.41] |

Source: Scottish Longitudinal Study
\* Number of deaths are calculated over 5 years post multimorbidity onset.

## Discussion

This study shows that it is possible to distinguish different typical sequences to multimorbidity based on a small number of diseases (diabetes, CVD and cancer). Using sequence and cluster analysis, we found seven typical trajectories to multimorbidity: later fast transition to multimorbidity, CVD start with slow transition to multimorbidity, cancer start with slow transition to multimorbidity, diabetes start with slow transition to multimorbidity, fast transition to both diabetes and CVD, fast transition to multimorbidity and death, fast transition to both cancer and CVD. We found sociodemographic differences between typical trajectories with individuals quickly transitioning to multimorbidity and death (cluster 6) showing the worse sociodemographic profile. In contrast, trajectories with a later multimorbidity start (cluster 1) or typical trajectories involving cancer (clusters 3 & 7) showed a relatively better socioeconomic profile compared to other typical multimorbidity trajectories. We also found some trajectories more common in males (cluster 2 - *CVD start, slow transition to multimorbidity*) or in females (cluster 3 - *cancer start, slow transition to multimorbidity*). In relation to hospitalisation outcomes post multimorbidity onset, individuals quickly transitioning to multimorbidity and death (cluster 6) showed significantly more hospitalisation and overnight stay relative to their exposure time (adjusted person-year) and after adjustment for sociodemographic determinants and other comorbidities. We also found significant differences in 5-year mortality outcome post multimorbidity onset, with trajectories of quick transition to multimorbidity and death (cluster 6) expectedly showing the highest risk of mortality, and trajectories of quick transition to both diabetes and CVD (cluster 5) and typical trajectories involving cancer (clusters 3 & 7) showing significantly lower 5-year mortality risk than the other typical trajectories.

Social science researchers have used single and multi-channel sequence analysis to understand life course processes (Studer and Ritschard, 2016). However, this study is the first to apply a single-channel sequence analysis with the aim to understand the sequencing of diseases leading to multimorbidity. Our analysis described typical multimorbidity trajectories, how they vary socio-demographically and are associated with different outcomes. These findings demonstrate the value of using sequence analysis in multimorbidity research, which represents one of the most challenging public health problems we face. More generally, we argue that multimorbidity research could benefit from taking an interdisciplinary approach and drawing methodological inspiration from non-medical disciplines to advance our understanding of multimorbidity development and associated risk factors.

A major aspect to consider in the application of sequence analysis to multimorbidity research is that identifying sequencing with precision relies on capturing onset of disease as accurately as possible. Such endeavour is challenging as administrative records depend on diagnosis and can only provide an approximation of onset. For example, a late diagnosis would not reflect the actual onset of disease. We choose commonly occurring diseases, where for example diagnosis for diabetes relies on a national diabetes register, including data from primary and secondary care (likewise for cancer). In the United Kingdom, there has been an increased incentive to improve care and reporting on chronic diseases such as diabetes, CVD and cancer under the Quality and Outcomes Framework introduced in 2004 and retired in 2016 (Kontopantelis et al., 2013; Roland and Guthrie, 2016). Hence, we have captured the order of diseases as accurately as possible based on the available Scottish administrative data. Any method researching multimorbidity and relying on onset identification will be faced by similar issues. One way to capture onset more accurately would be to prospectively follow individuals and continuously monitor their health for the identification of a specific list of diseases. Such targeted health follow-up might be possible in a longitudinal prospective survey albeit based on smaller sample size than what administrative data can provide. In our application of sequence analysis to the study of multimorbidity trajectory, disease can only accumulate. For example, once an individual has been diagnosed with cancer, this accumulates with other diseases even though a complete remission may have occurred. Information on remission was not available from our data. This has implications on the interpretation of our findings. For example, we found that the typical trajectory involving a cancer start (cluster 3) had a statistically significant lower risk of mortality than typical trajectories involving a CVD or diabetes start (clusters 2 and 4). One explanation could be that this typical multimorbidity trajectory with a cancer start might be dominated by cancer survivors who are relatively healthy with slow onset of other diseases. Our analysis was based on sequences reconstructed from three diseases only and this resulted in a diversity of typical trajectories highlighting the complexity of multimorbidity trajectories more generally. Indeed, sequence analysis can be useful to disentangle the complex pathways to multimorbidity based on a limited set of diseases. However, an analysis based on four or five diseases would produce respectively 16 or 32 states and a sequence analysis based on more than 12 states becomes very difficult to interpret, a recognised limitation of this methodology. Therefore, it is not practical nor interpretable to use four or more diseases with the approach described in this paper. Consequently, sequence analysis in the context of exploring disease trajectories fits better with a clearly defined and theoretically driven focus (e.g. two or three diseases). This is not necessarily a limitation in specialist clinical settings where disease clustering typically involves a relatively small number of conditions. With a limited number of diseases of interest and a more detailed data on disease progression, multi-channel sequence analysis could also be an option taking one disease per channel and exploring multiple disease trajectories concomitantly.

To identify typical trajectories using a sequence analysis approach, a series of choices at OM and cluster analysis stages must be made. This is a known limitation of this methodology (Abbott and Tsay, 2000; Brzinsky-Fay and Kohler, 2010; Studer and Ritschard, 2016). The choice of dissimilarity costs and clustering methods have an influence on the number of clusters and the content of the final clusters. As a robustness check, it is advised to vary specifications and assess whether different specifications alter findings. We tested different specifications and clustering quality measures of some of these sensitivity analyses are available in Additional file 1. Clusters resulting from sensitivity analyses showed similar typical multimorbidity trajectories than those presented here.

The analysis of multiple diseases longitudinally remains a challenge, especially if one would rather capture the specificity of each disease in the process of disease accumulation than just count diseases. Data mining or machine learning approaches can cater for a higher number of chronic diseases to explore multimorbidity trajectories, but it can be challenging to interpret the multitude of multimorbidity trajectories identified. This remains a real issue if we wish to extract meaningful information for practice and health services, particularly in generalist settings. Focusing on a specific set of diseases for extracting the drivers of complex disease trajectories has the potential to inform policy and health services on prevention and tailored interventions for a targeted set of trajectories.

## Conclusions

Understanding the sociodemographic profile and outcomes of diverse typical trajectories of multimorbidity is complex. This complexity was revealed using sequence analysis based on a small set of commonly occurring diseases (diabetes, CVD and cancer) to characterise trajectories to multimorbidity. Further methodological work should consider scaling up to more diseases, or if disease progression data were available, using multi-channel sequence analysis to explore parallel detailed disease trajectories. However, when considering more than a few diseases, other methods, such as machine learning, may be more appropriate. Scaling up, however, will come with the challenge of interpreting findings and drawing meaningful conclusions from a very large number of trajectories.

## Data Availability

Data is available through application and relevant approval from the Scottish Longitudinal Study (SLS).

## List of abbreviations

CI: confidence intervals
CVD: cardiovascular disease
HR: hazard ratio
ICD10: International Classification of Disease version 10
LSCS: Longitudinal Studies Centre – Scotland
OM: optimal matching
OR: odd ratio
RR: risk ratio
SIMD: Scottish Index for Multiple Deprivation
SLS: Scottish Longitudinal Study
SM: substitution matrix

## Declarations

### Ethics approval and consent to participate

We obtained ethical approval for this study from the University Teaching and Research Ethics Committee at the University of St Andrews (reference GG14300). Our SLS study linking the Scottish censuses to health record was approved by the SLS Research Board (SLS project number 2018_012) and the Public Benefit and Privacy Panel for Health and Social Care of NHS Scotland in October 2019 (reference 1819-0093). Individual consent was not sought. SLS linked datasets are anonymised and available in a dedicated safe haven following a strict protocol on access and disclosure control to ensure the safety and confidentiality of the data. Access is restricted to named researchers with appropriate training and clearance.

### Consent for publication

Not applicable.

### Availability of data and materials

The data that support the findings of this study are available from SLS but restrictions apply to the availability of these data, which were used under license for the current study, and so are not publicly available. Data are however available from the authors upon reasonable request and with permission of SLS.

### Authors’ Contributions

KK and GC conceptualised and refined the idea for the study helped by FS. GC analysed the data and wrote the manuscript. KK acquired the funding and helped draft the manuscript. FS provided a clinical perspective to refine the direction of the study. All authors read and discussed all drafts of the manuscript and approved the final version for submission.

## Acknowledgements

We thank Julia Mikolai for her methodological support and feedbacks. Prof Colin McCowan supported our approach to longitudinal multimorbidity research by providing access to a pilot multimorbidity study within the Health Informatics Centre (HIC) for methodological testing purpose. The help provided by staff of the Longitudinal Studies Centre – Scotland (LSCS) is also acknowledged. The LSCS is supported by the Economic and Social Research Council and Joint Information Systems Committee census programme, the Scottish Funding Council, the Chief Scientist’s Office and the Scottish Government. The authors alone are responsible for the interpretation of the data. Census output is Crown copyright and is reproduced with the permission of the Controller of Her Majesty’s Stationery Office and the Queen’s Printer for Scotland.

