## Additional file 1 for "Understanding multimorbidity trajectories in Scotland: an application of sequence analysis"

**Additional file 1. Cluster quality measures for different specifications of the indel value**

**C**luster quality measures allow the identification of the best clustering solution. A range of measures are available in R using the wcKMedRange function from the WeightedCluster library: PBC - Point Biserial Correlation, HG - Hubert's Gamma, HGSD - Hubert's Somers D, ASW - the average value of the silhouette, ASWw - the average value of the silhouette weighted, CH - Calinski Harabasz Index, CHsq - CH squared, R2 – Pseudo R2, R2sq – R2 squared, HC - Hubert's C.

For interpretation, an ASW measure close to 1 means the corresponding n-clusters solution is well clustered. A HC close to 0 means good clustering. For other measures (PBC, HG, HGSD, CH, CHsq, R2, R2sq), higher values mean better clustering.

See below the cluster quality measures obtained for the sensitivity analyses using varied Indel values: 0.5, 1, 1.5 and 5.

a. Indel value of 0.5

PBC	HG HGSD ASW ASWw	CH	R2	CHsq	R2sq	HC

cluster2	0.22	0.27	0.26	0.19	0.19	579.61	0.11	927.50	0.16	0.32

cluster2	0.30	0.37	0.35	0.20	0.20	857.88	0.12	1415.50	0.18	0.29

cluster3	0.39	0.44	0.42	0.21	0.21	839.68	0.21	1441.17	0.31	0.30

cluster4	0.50	0.60	0.59	0.25	0.25	854.63	0.29	1587.74	0.43	0.25

cluster5	0.52	0.64	0.63	0.28	0.28	867.15	0.36	1706.25	0.52	0.23

cluster6	0.59	0.77	0.76	0.29	0.29	891.64	0.41	1925.04	0.60	0.17

cluster7	0.63	0.85	0.85	0.32	0.32	876.36	0.46	2069.86	0.66	0.12

cluster8	0.64	0.87	0.87	0.32	0.32	821.35	0.48	2033.51	0.69	0.11

cluster9	0.61	0.87	0.87	0.29	0.29	781.16	0.50	1966.13	0.71	0.12

cluster10	0.57	0.87	0.86	0.26	0.26	749.67	0.52	1900.78	0.73	0.13

cluster11	0.55	0.87	0.87	0.25	0.26	718.48	0.53	1835.90	0.74	0.13

cluster12	0.54	0.87	0.86	0.25	0.25	691.64	0.55	1757.47	0.75	0.13

The cluster quality measures above point to a good clustering for a 7 or 8-clusters solution.

b. Indel value of 1

PBC	HG HGSD ASW ASWw	CH	R2	CHsq	R2sq	HC

cluster2	0.28	0.32	0.30	0.18	0.18	708.33	0.10	1208.42	0.16	0.29

cluster3	0.44	0.53	0.51	0.21	0.21	701.68	0.18	1227.04	0.28	0.28

cluster4	0.46	0.55	0.53	0.21	0.21	726.84	0.26	1296.62	0.38	0.29

cluster5	0.54	0.69	0.68	0.24	0.24	753.78	0.32	1461.07	0.48	0.23

cluster6	0.55	0.72	0.71	0.26	0.26	769.04	0.38	1550.72	0.55	0.22

cluster7	0.58	0.76	0.75	0.27	0.27	725.08	0.41	1534.57	0.59	0.20

cluster8	0.56	0.78	0.77	0.25	0.25	688.42	0.43	1476.17	0.62	0.19

cluster9	0.57	0.81	0.81	0.25	0.25	656.75	0.45	1436.61	0.65	0.17

cluster10	0.57	0.83	0.83	0.26	0.26	635.28	0.48	1458.59	0.68	0.16

cluster11	0.55	0.83	0.82	0.25	0.25	621.47	0.50	1433.91	0.70	0.16

cluster12	0.55	0.83	0.83	0.25	0.25	608.45	0.52	1429.10	0.71	0.16

The cluster quality measures above point to a good clustering for a 6 or 7-clusters solution.

c. Indel value of 1.5

PBC	HG HGSD ASW ASWw	CH	R2	CHsq	R2sq	HC

cluster2	0.20	0.19	0.18	0.15	0.15	715.86	0.10	1160.98	0.16	0.34

cluster3	0.40	0.46	0.44	0.19	0.19	707.58	0.18	1212.64	0.28	0.31

cluster4	0.42	0.48	0.46	0.21	0.21	718.62	0.25	1252.24	0.37	0.31

cluster5	0.54	0.68	0.66	0.24	0.24	728.95	0.32	1390.58	0.47	0.23

cluster6	0.59	0.76	0.75	0.26	0.26	726.03	0.37	1472.11	0.54	0.20

cluster7	0.60	0.77	0.77	0.26	0.27	693.23	0.40	1467.16	0.58	0.19

cluster8	0.57	0.77	0.76	0.25	0.25	667.18	0.43	1430.63	0.61	0.20

cluster9	0.59	0.82	0.82	0.25	0.25	645.86	0.45	1443.59	0.65	0.17

cluster10	0.58	0.84	0.83	0.26	0.26	631.20	0.47	1451.06	0.67	0.16

cluster11	0.56	0.85	0.84	0.26	0.26	619.54	0.50	1450.08	0.70	0.16

cluster12	0.57	0.87	0.87	0.27	0.27	604.00	0.51	1497.33	0.72	0.14

The cluster quality measures above point to a good clustering for a 7-clusters solution.

d. Indel value of 5

PBC	HG HGSD ASW ASWw	CH	R2	CHsq	R2sq	HC

cluster2	0.20	0.19	0.18	0.15	0.15	716.10	0.10	1161.31	0.16	0.34

cluster3	0.40	0.46	0.44	0.19	0.19	707.81	0.18	1213.05	0.28	0.31

cluster4	0.42	0.48	0.46	0.21	0.21	718.80	0.26	1252.60	0.37	0.31

cluster5	0.54	0.68	0.66	0.24	0.24	729.19	0.32	1391.17	0.47	0.23

cluster6	0.59	0.76	0.75	0.26	0.26	726.29	0.37	1472.87	0.54	0.20

cluster7	0.60	0.77	0.77	0.26	0.27	693.47	0.40	1467.94	0.58	0.19

cluster8	0.57	0.77	0.76	0.25	0.25	667.41	0.43	1431.37	0.61	0.20

cluster9	0.59	0.82	0.82	0.25	0.25	645.01	0.45	1439.90	0.65	0.17

cluster10	0.58	0.84	0.83	0.26	0.26	630.38	0.47	1447.20	0.67	0.16

cluster11	0.56	0.84	0.84	0.26	0.26	618.60	0.50	1445.43	0.70	0.16

cluster12	0.57	0.87	0.87	0.27	0.27	602.56	0.51	1489.51	0.72	0.14

The cluster quality measures above provide very similar results to that of the analysis with an indel of 1.5 and point to a good clustering for a 7-clusters solution.
