## Additional file 2 for "Understanding multimorbidity trajectories in Scotland: an application of sequence analysis"

**Additional file 2. Sex and age-adjusted multinomial logistic regressions.**

|  |  |  | **Model with age and sex** | **Model with age, sex, and marital status** | **Model with age, sex, and household size** |
| --- | --- | --- | --- | --- | --- |
| **Covariate** | **Category** | **Cluster**  (cluster 6 as reference)* | **OR [95% CI]** | **OR [95% CI]** | **OR [95% CI]** |
| Age (continuous) |  | 1 | 0.94 [0.93,0.95] | 0.94 [0.93,0.95] | 0.94 [0.93,0.95] |
|  |  | 2 | 0.95 [0.94,0.96] | 0.95 [0.94,0.96] | 0.95 [0.94,0.96] |
|  |  | 3 | 0.96 [0.94,0.97] | 0.96 [0.94,0.97] | 0.96 [0.94,0.97] |
|  |  | 4 | 0.94 [0.93,0.96] | 0.94 [0.93,0.96] | 0.95 [0.93,0.96] |
|  |  | 5 | 0.93 [0.92,0.94] | 0.93 [0.92,0.94] | 0.94 [0.92,0.95] |
|  |  | 7 | 0.97 [0.96,0.99] | 0.98 [0.96,0.99] | 0.97 [0.96,0.99] |
| Sex | Male vs Female | 1 | 0.96 [0.82,1.23] | 0.93 [0.79,1.09] | 0.93 [0.79,1.09] |
|  |  | 2 | 1.28 [1.06,1.54] | 1.24 [1.02,1.50] | 1.24 [1.02,1.49] |
|  |  | 3 | 0.62 [0.48,0.79] | 0.57 [0.45,0.74] | 0.59 [0.46,0.76] |
|  |  | 4 | 1.01 [0.80,1.28] | 0.98 [0.78,1.25] | 1.00 [0.79,1.27] |
|  |  | 5 | 0.97 [0.79,1.19] | 0.92 [0.74,1.13] | 0.93 [0.76,1.15] |
|  |  | 7 | 0.85 [0.66,1.09] | 0.79 [0.61,1.02] | 0.81 [0.63,1.04] |
| Marital status | Married vs Single | 1 |  | 1.29 [0.95,1.75] |  |
|  |  | 2 |  | 1.56 [1.09,2.24] |  |
|  |  | 3 |  | 2.20 [1.26,3.84] |  |
|  |  | 4 |  | 1.10 [0.72,1.69] |  |
|  |  | 5 |  | 1.52 [1.01,2.27] |  |
|  |  | 7 |  | 2.09 [1.18,3.71] |  |
|  | Separated/Divorced/Widowed | 1 |  | 1.08 [0.78,1.50] |  |
|  | vs Single | 2 |  | 1.31 [0.89,1.93] |  |
|  |  | 3 |  | 1.47 [0.81,2.66] |  |
|  |  | 4 |  | 0.98 [0.61,1.55] |  |
|  |  | 5 |  | 1.11 [0.72,1.72] |  |
|  |  | 7 |  | 1.39 [0.75,2.57] |  |
| Household size | Household with 2 people | 1 |  |  | 1.19 [0.98,1.45] |
|  | vs 1-person | 2 |  |  | 1.23 [0.98,1.54] |
|  |  | 3 |  |  | 1.44 [1.06,1.96] |
|  |  | 4 |  |  | 0.93 [0.70,1.24] |
|  |  | 5 |  |  | 1.27 [0.98,1.66] |
|  |  | 7 |  |  | 1.51 [1.10,2.08] |
|  | Household with 3 or more people | 1 |  |  | 1.31 [1.02,1.87] |
|  | vs 1-person | 2 |  |  | 1.42 [1.07,1.90] |
|  |  | 3 |  |  | 1.06 [0.71,1.58] |
|  |  | 4 |  |  | 1.18 [0.84,1.67] |
|  |  | 5 |  |  | 1.59 [1.15,2.18] |
|  |  | 7 |  |  | 1.16 0.76,1.77] |

Source: Scottish Longitudinal Study

* Each cluster represents a typical multimorbidity trajectory as follows: cluster 1 - later fast transition to multimorbidity; cluster 2 - CVD start with slower transition to multimorbidity; cluster 3 - cancer start with slower transition to multimorbidity; cluster 4 - diabetes start with slower transition to multimorbidity; cluster 5 - fast transition to both diabetes and CVD; cluster 6 - fast transition to multimorbidity and death; cluster 7 - fast transition to both cancer and CVD.

|  |  |  | **Model with age, sex, and education** | **Model with age, sex, and household tenure** | **Model with age, sex, and SIMD** |
| --- | --- | --- | --- | --- | --- |
| **Covariate** | **Category** | **Cluster**  (cluster 6 as reference)* | **OR [95% CI]** | **OR [95% CI]** | **OR [95% CI]** |
| Age (continuous) |  | 1 | 0.94 [0.93,0.95] | 0.94 [0.93,0.95] | 0.94 [0.93,0.95] |
|  |  | 2 | 0.95 [0.94,0.96] | 0.95 [0.94,0.96] | 0.95 [0.94,0.96] |
|  |  | 3 | 0.96 [0.95,0.98] | 0.96 [0.94,0.97] | 0.96 [0.94,0.97] |
|  |  | 4 | 0.94 [0.93,0.95] | 0.94 [0.93,0.95] | 0.95 [0.93,0.96] |
|  |  | 5 | 0.93 [0.92,0.94] | 0.93 [0.92,0.94] | 0.94 [0.92,0.95] |
|  |  | 7 | 0.98 [0.96,0.99] | 0.97 [0.96,0.99] | 0.97 [0.96,0.99] |
| Sex | Male vs Female | 1 | 0.94 [0.80,1.11] | 0.93 [0.79,1.10] | 0.93 [0.79,1.09] |
|  |  | 2 | 1.28 [1.06,1.54] | 1.27 [1.06,1.53] | 1.24 [1.02,1.49] |
|  |  | 3 | 0.59 [0.46,0.76] | 0.59 [0.46,0.76] | 0.59 [0.46,0.76] |
|  |  | 4 | 1.01 [0.80,1.27] | 0.99 [0.78,1.25] | 1.00 [0.79,1.27] |
|  |  | 5 | 0.96 [0.78,1.18] | 0.96 [0.78,1.18] | 0.93 [0.76,1.15] |
|  |  | 7 | 0.83 [0.64,1.06] | 0.81 [0.63,1.05] | 0.81 [0.63,1.04] |
| Educational levels | High educational level | 1 | 1.44 [1.13,1.84] |  |  |
|  | vs No qualification | 2 | 1.08 [0.81,1.43] |  |  |
|  |  | 3 | 2.07 [1.47,2.91] |  |  |
|  |  | 4 | 1.28 [0.91,1.80] |  |  |
|  |  | 5 | 1.12 [0.81,1.53] |  |  |
|  |  | 7 | 1.60 [1.11,2.29] |  |  |
|  | Low educational level | 1 | 1.05 [0.86,1.28] |  |  |
|  | vs No qualification | 2 | 0.90 [0.71,1.13] |  |  |
|  |  | 3 | 1.24 [0.91,1.68] |  |  |
|  |  | 4 | 0.80 [0.59,1.09] |  |  |
|  |  | 5 | 1.05 [0.82,1.36] |  |  |
|  |  | 7 | 1.05 [0.76,1.45] |  |  |
| Household tenure | Owned vs social rented | 1 |  | 1.45 [1.22,1.73] |  |
|  |  | 2 |  | 1.16 [0.95,1.41] |  |
|  |  | 3 |  | 1.79 [1.35,2.38] |  |
|  |  | 4 |  | 1.40 [1.08,1.81] |  |
|  |  | 5 |  | 1.12 [0.90,1.40] |  |
|  |  | 7 |  | 1.66 [1.25,2.22] |  |
|  | Private rented vs social rented | 1 |  | 1.35 [0.83,2.21] |  |
|  |  | 2 |  | 0.80 [0.44,1.46] |  |
|  |  | 3 |  | 0.71 [0.26,1.90] |  |
|  |  | 4 |  | 1.05 [0.50,2.20] |  |
|  |  | 5 |  | 0.92 [0.49,1.75] |  |
|  |  | 7 |  | 0.90 [0.36,2.26] |  |
|  | Live rent-free vs social rented | 1 |  | 1.09 [0.66,1.79] |  |
|  |  | 2 |  | 1.52 [0.91,2.54] |  |
|  |  | 3 |  | 1.62 [0.79,3.31] |  |
|  |  | 4 |  | 1.25 [0.62,2.54] |  |
|  |  | 5 |  | 0.92 [0.48,1.76] |  |
|  |  | 7 |  | 1.13 [0.50,2.57] |  |
| SIMD (continuous) |  | 1 |  |  | 1.10 [1.04,1.17] |
|  |  | 2 |  |  | 1.01 [0.94,1.08] |
|  |  | 3 |  |  | 1.24 [1.13,1.35] |
|  |  | 4 |  |  | 1.05 [0.97,1.15] |
|  |  | 5 |  |  | 1.03 [0.96,1.11] |
|  |  | 7 |  |  | 1.15 [1.05,1.25] |

Source: Scottish Longitudinal Study

* Each cluster represents a typical multimorbidity trajectory as follows: cluster 1 - later fast transition to multimorbidity; cluster 2 - CVD start with slower transition to multimorbidity; cluster 3 - cancer start with slower transition to multimorbidity; cluster 4 - diabetes start with slower transition to multimorbidity; cluster 5 - fast transition to both diabetes and CVD; cluster 6 - fast transition to multimorbidity and death; cluster 7 - fast transition to both cancer and CVD.
