## Additional file 3 for "Understanding multimorbidity trajectories in Scotland: an application of sequence analysis"

**Additional file 3. Fully adjusted multinomial logistic regressions.**

|  |  |  | **Model with age, sex, marital status, household size, educational level, household tenure and SIMD** |
| --- | --- | --- | --- |
| **Covariate** | **Category** | **Cluster**  (cluster 6 as reference)* | **OR [95% CI]** |
| Age (continuous) |  | 1 | 0.94 [0.93,0.95] |
|  |  | 2 | 0.95 [0.94,0.96] |
|  |  | 3 | 0.95 [0.94,0.97] |
|  |  | 4 | 0.94 [0.93,0.96] |
|  |  | 5 | 0.94 [0.92,0.95] |
|  |  | 7 | 0.97 [0.95,0.99] |
| Sex | Male vs Female | 1 | 0.91 [0.77,1.07] |
|  |  | 2 | 1.24 [1.02,1.50] |
|  |  | 3 | 0.56 [0.43,0.72] |
|  |  | 4 | 0.97 [0.77,1.24] |
|  |  | 5 | 0.91 [0.74,1.13] |
|  |  | 7 | 0.77 [0.60,1.00] |
| Marital status | Married vs Single | 1 | 1.15 [0.80,1.65] |
|  |  | 2 | 1.47 [0.97,2.25] |
|  |  | 3 | 2.48 [1.28,4.83] |
|  |  | 4 | 1.23 [0.73,2.07] |
|  |  | 5 | 1.46 [0.91,2.34] |
|  |  | 7 | 2.13 [1.08,4.18] |
|  | Separated/Divorced/Widowed | 1 | 1.08 [0.78,1.51] |
|  | vs Single | 2 | 1.31 [0.89,1.94] |
|  |  | 3 | 1.51 [0.83,2.76] |
|  |  | 4 | 0.96 [0.60,1.53] |
|  |  | 5 | 1.09 [0.70,1.69] |
|  |  | 7 | 1.42 [0.77,2.62] |
| Household size | Household with 2 people | 1 | 1.03 [0.76,1.39] |
|  | vs 1-person | 2 | 1.05 [0.75,1.49] |
|  |  | 3 | 0.77 [0.46,1.29] |
|  |  | 4 | 0.70 [0.45,1.09] |
|  |  | 5 | 0.97 [0.65,1.44] |
|  |  | 7 | 0.89 [0.52,1.51] |
|  | Household with 3 or more people | 1 | 1.16 [0.83,1.62] |
|  | vs 1-person | 2 | 1.23 [0.84,1.81] |
|  |  | 3 | 0.58 [0.33,1.04] |
|  |  | 4 | 0.91 [0.56,1.48] |
|  |  | 5 | 1.23 [0.79,1.89] |
|  |  | 7 | 0.70 [0.39,1.27] |
| Educational levels | High educational level | 1 | 1.26 [0.98,1.63] |
|  | vs No qualification | 2 | 1.05 [0.78,1.42] |
|  |  | 3 | 1.60 [1.11,2.29] |
|  |  | 4 | 1.14 [0.80,1.64] |
|  |  | 5 | 1.09 [0.78,1.52] |
|  |  | 7 | 1.31 [0.89,1.92] |
|  | Low educational level | 1 | 0.97 [0.78,1.19] |
|  | vs No qualification | 2 | 0.88 [0.69,1.11] |
|  |  | 3 | 1.06 [0.77,1.45] |
|  |  | 4 | 0.75 [0.55,1.02] |
|  |  | 5 | 1.03 [0.79,1.33] |
|  |  | 7 | 0.92 [0.66,1.29] |
| Household tenure | Owned vs social rented | 1 | 1.31 [1.08,1.59] |
|  |  | 2 | 1.12 [0.90,1.40] |
|  |  | 3 | 1.32 [0.97,1.81] |
|  |  | 4 | 1.40 [1.05,1.87] |
|  |  | 5 | 1.01 [0.79,1.29] |
|  |  | 7 | 1.39 [1.01,1.90] |
|  | Private rented vs social rented | 1 | 1.27 [0.77,2.08] |
|  |  | 2 | 0.82 [0.45,1.49] |
|  |  | 3 | 0.57 [0.21,1.55] |
|  |  | 4 | 1.03 [0.49,2.17] |
|  |  | 5 | 0.91 [0.47,1.73] |
|  |  | 7 | 0.81 [0.32,2.05] |
|  | Live rent-free vs social rented | 1 | 1.08 [0.66,1.78] |
|  |  | 2 | 1.58 [0.94,2.65] |
|  |  | 3 | 1.54 [0.75,3.15] |
|  |  | 4 | 1.25 [0.61,2.53] |
|  |  | 5 | 0.96 [0.50,1.84] |
|  |  | 7 | 1.13 [0.50,2.56] |
| SIMD (continuous) |  | 1 | 1.05 [0.98,1.12] |
|  |  | 2 | 0.99 [0.92,1.06] |
|  |  | 3 | 1.14 [1.03,1.26] |
|  |  | 4 | 1.01 [0.92,1.12] |
|  |  | 5 | 1.01 [0.93,1.10] |
|  |  | 7 | 1.07 [0.96,1.18] |

Source: Scottish Longitudinal Study

* Each cluster represents a typical multimorbidity trajectory as follows: cluster 1 - later fast transition to multimorbidity; cluster 2 - CVD start with slower transition to multimorbidity; cluster 3 - cancer start with slower transition to multimorbidity; cluster 4 - diabetes start with slower transition to multimorbidity; cluster 5 - fast transition to both diabetes and CVD; cluster 6 - fast transition to multimorbidity and death; cluster 7 - fast transition to both cancer and CVD.
